# Pathomic Texture Composites from H&E, and Immunohistochemistry of Prostatectomy Specimens for Exploratory Modeling of Time to Biochemical Recurrence

**DOI:** 10.64898/2026.10.23.26366338

**Authors:** Oluwaseyi M Oderinde, Ruchika Reddy Chimmula, Varsha Nair, Jordan Holmes, Clinton Bahler

## Abstract

**Background:** Pathomic analysis, the extraction of quantitative features from digitized histology, can capture tissue patterns that may complement conventional prognostic factors. We explored whether stain-specific texture composites derived from hematoxylin and eosin (H&E), CD8, and CD163 immunohistochemistry add information to models of time to biochemical recurrence (BCR) after radical prostatectomy.

**Methods:** This retrospective exploratory study included 15 patients with digitized prostatectomy specimens and linked clinicopathologic data. Handcrafted pathomic features were extracted from image tiles with PyRadiomics, averaged within patients, and standardized. Three stain-specific, equal-weight texture composites developed for this study were integrated with clinicopathologic variables in Cox models with an L2 penalty of 0.1. Four predictor configurations were compared using apparent concordance indices (C-indices). Fitted survival functions were used to explore recurrence probabilities over six-month intervals.

**Results:** Apparent C-indices were 0.66 (95% CI, 0.38–0.85) for clinicopathologic variables alone, 0.68 (0.38– 0.86) after adding H&E, 0.88 (0.75–0.95) after adding CD8 and CD163, and 0.89 (0.76–0.96) with all three composites. The increase was confined to the immune-stain composites. Composite log-hazard coefficients were approximately 1.06 for CD8, 0.51 for CD163, and −0.10 for H&E.

**Conclusion:** Stain-specific pathomic composites are a compact way to integrate immunohistochemical and H&E texture with clinicopathologic predictors. The gain in apparent discrimination with CD8 and CD163 texture is hypothesis-generating; larger, representative cohorts with independent validation are needed to establish incremental prognostic value and reliable absolute recurrence probabilities.

## 1. Introduction

Following radical prostatectomy, prostate-specific antigen (PSA) measurements provide a reproducible basis for identifying biochemical recurrence (BCR), provided the PSA threshold and confirmation rule are specified. (Cookson et al., 2007). The subsequent clinical course is heterogeneous: BCR, metastatic progression, and prostate cancer death are distinct outcomes, and the intervals between them vary substantially. (Freedland et al., 2005; Pound, 1999) A model of time to first postoperative BCR therefore addresses an earlier prognostic question than a model of metastasis or mortality after recurrence.

Established postoperative tools combine preoperative PSA with surgical pathology to estimate recurrence risk. CAPRA-S and postoperative nomograms illustrate the information contained in grade, stage, surgical margins, extraprostatic extension, seminal vesicle invasion, and nodal involvement. (Cooperberg et al., 2011; Stephenson et al., 2005). The ISUP Grade Group framework and prognostically relevant architectural patterns further organize the information available from routine pathology (Epstein et al., 2016). In Gleason score 7 disease, for example, cribriform growth has been associated with metastasis and disease-specific death (Kweldam et al., 2015). Genomic classifiers provide an additional benchmark for evaluating whether a new tissue-derived measurement offers information beyond conventional clinicopathologic assessment (Erho et al., 2013). New image-derived markers must establish what they add to these existing assessments.

Digital pathology enables quantitative analysis of the same specimens used for routine diagnosis. Open whole-slide analysis platforms support tissue annotation and measurement, (Leo et al., 2021) while computational studies have related gland morphology and tissue-structure features to BCR. (Leo et al., 2021; Marin et al., 2025) These findings motivate analysis of image patterns that are difficult to summarize by visual inspection alone. Feature extractors developed for radiomics, such as PyRadiomics, provide mathematically defined intensity and texture measurements that can also be computed on digitized histology, where they are termed pathomic features. Their reproducibility, however, depends on image preparation and acquisition as well as the feature definitions. (Van Griethuysen et al., 2017; Zwanenburg et al., 2020)

Immune staining supplies complementary information about the prostate tumor microenvironment. Higher epithelial CD8-positive lymphocyte density has been associated with shorter biochemical failure-free survival in a prostatectomy cohort, demonstrating that marker abundance does not have a uniformly favorable prognostic interpretation. (Ness et al., 2014) Separate studies have associated infiltrating regulatory T cells and macrophages with BCR, and peritumoral CD163-positive macrophages with late adverse outcomes. (Andersen et al., 2021; Molina et al., 2023) These observations support examining immune-staining patterns while distinguishing image texture, cell abundance, and cellular function.

The specific question addressed here is whether a small set of stain-specific composites can condense pathomic measurements from H&E, CD8, and CD163 images into useful inputs for postoperative time-to-event modeling. This approach does not require explicit enumeration of cells or estimation of epithelial, stromal, and luminal proportions. It may therefore capture complementary aspects of local staining heterogeneity with fewer segmentation requirements. We developed three equal-weight composites, integrated them with clinicopathologic variables in penalized Cox models, and assessed measurement feasibility and apparent discrimination to inform a subsequent validation study.

## 2. Materials and Methods

### 2.1 Study design and clinical endpoint

We conducted a retrospective exploratory analysis of 15 patients with prostate cancer, available radical prostatectomy specimens, and linked clinicopathologic and follow-up data. Each patient was an independent observational unit; multiple sections and image tiles contributed to measuring each patient’s features. The intended application was postoperative risk assessment using information available from the surgical specimen and clinical record.

The endpoint was PSA-defined BCR, with time expressed in months from definitive treatment to recorded recurrence or last follow-up. The intended censoring rule was the last recurrence-free assessment for patients without an observed event. A PSA threshold and confirmation rule are essential to make this endpoint reproducible; postoperative PSA persistence also needs to be distinguished from recurrence after an initially undetectable PSA. The commonly used research definition of a PSA of at least 0.2 ng/mL followed by confirmatory elevation provides a reporting reference (Cookson et al., 2007) rather than evidence that this exact rule was implemented in the present cohort.

Candidate clinical variables include baseline prostate-specific antigen (PSA), Gleason score from biopsy, extraprostatic extension (EPE), seminal vesicle invasion (SV), surgical margin status, tumor stage, age, and time-to-event data (**Table 1**).

**Table 1.** Patient Characteristics.

| Variable | Description | Value (Example Summary) |
| --- | --- | --- |
| Number of patients | Total cohort size | 15 |
| Age (years) | Median (range) | ~65 (53–76) |
| PSA (baseline) | Median (range) | ~6–22 |
| Gleason Score | Biopsy-based grading | 1–4 (categorical) |
| EPE | Extraprostatic extension | 0/1/2 |
| SV | Seminal vesicle invasion | 0/1/2 |
| Margin | Surgical margin status | 0/1 |
| T stage | Tumor stage | 2–4 |
| Recurrence events | Number of BCR cases | 14 |
| Censored cases | Non-recurrence | 1 |
| Follow-up time (months) | Range | ~4–96 |

### 2.2 Tissue preparation and pathomic feature extraction

An experienced pathologist selected tumor-bearing tissue from the region with the largest tumor burden. Four representative tissue sections were described for each patient, with H&E, CD8, and CD163 staining yielding 12 whole-slide images (WSI) per patient. H&E provided architectural context, while the immunostains enabled analysis of the appearance and distribution of marker-associated staining. Sampling the dominant tumor region aimed to improve consistency of the analytic target, although it did not ensure representation of all tumor foci or spatial compartments.

All stained slides were digitized as high-resolution whole-slide images (WSIs). Each WSI was subsequently partitioned into non-overlapping image tiles after excluding background and non-tissue regions. This tiling strategy enabled localized quantitative analysis while preserving spatial information across the tissue.

Quantitative pathomic features describing tissue morphology, texture, intensity, and spatial heterogeneity were extracted from every tile independently using PyRadiomic library that was adapted for 2D digital pathology. These tile-level features served as the fundamental units for downstream feature engineering and were subsequently aggregated to generate patient-level biomarkers used for modeling in this study. The workflow is summarized in **Figure 1**.

**Figure 1.**
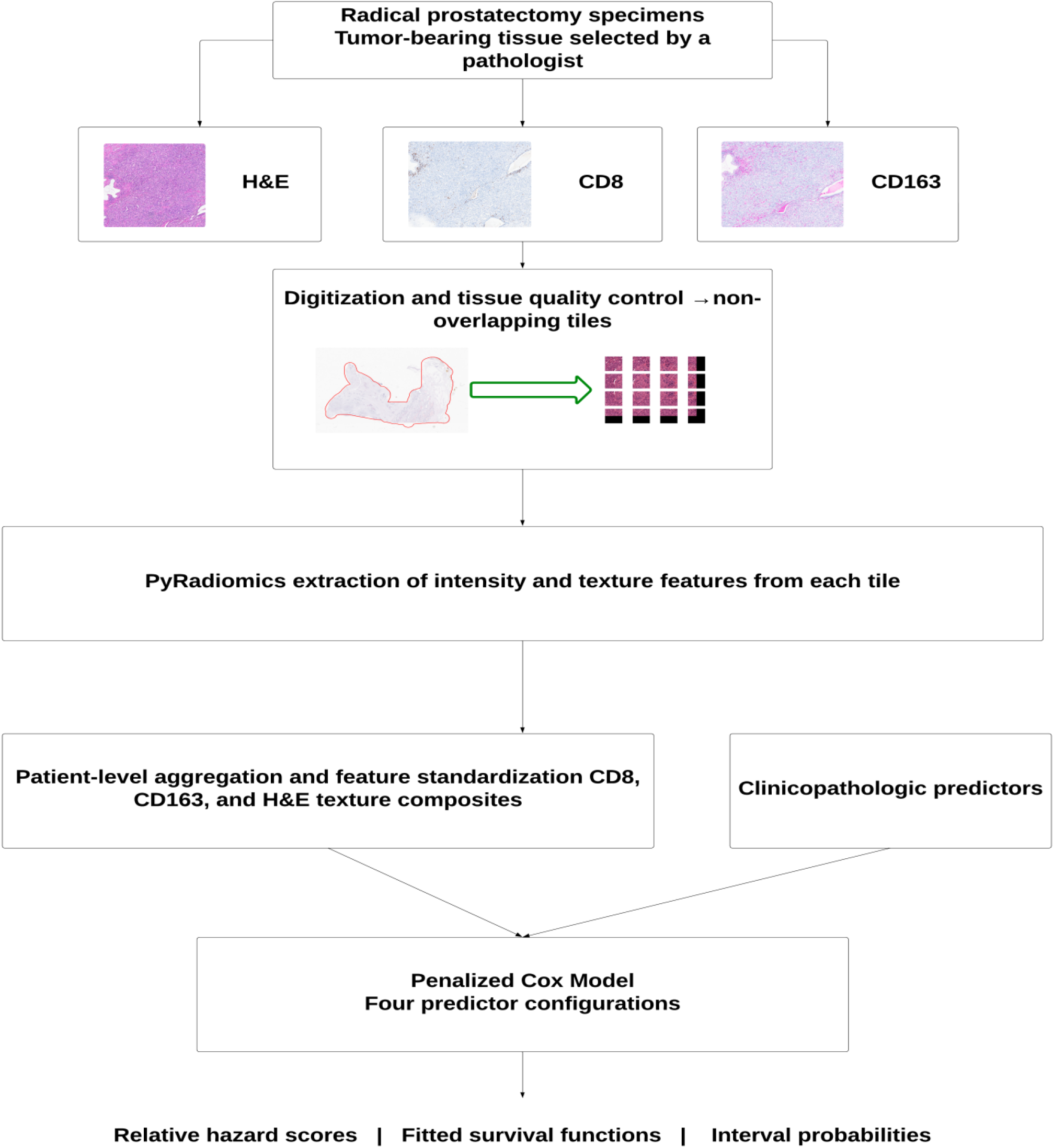
Analysis workflow. Pathomic measurements from H&E, CD8, and CD163 tiles are aggregated within patients, standardized, and combined into three texture composites. The composites and clinicopathologic predictors enter four penalized Cox-model configurations. Outputs include relative hazard scores, fitted BCR-free survival functions, and interval probabilities.

### 2.3 Patient-level aggregation and composite construction

Each aggregated feature was standardized using its across-patient mean and standard deviation. For patient i and feature j, the standardized value was defined as follows.

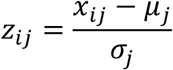

Here, x is the patient-level feature; μ is the feature’s mean, and σ is its standard deviation in the data used to estimate preprocessing parameters. In any resampled or external evaluation, these parameters must be estimated in the training data and applied unchanged to held-out patients; the same principle applies to imputation and any outcome-guided feature selection. Reporting should make this sequence explicit (Collins et al., 2024; McShane et al., 2005).

Three equal-weight composites were defined from standardized feature components. We use the neutral terms CD8 texture composite, CD163 texture composite, and H&E texture composite because the measurements have not been validated as assays of cellular activation, suppression, or morphologic disorder. Their algebraic definitions are retained below.

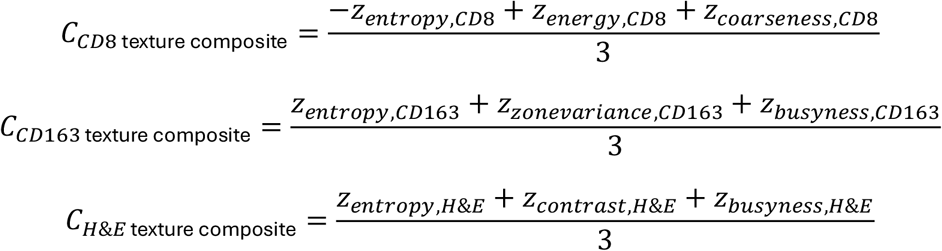

The z terms denote stain-specific standardized entropy, energy, coarseness, zone variance, busyness, or contrast. Exact feature-family definitions are necessary because names such as entropy and energy can refer to different mathematical quantities in different extraction implementations. Published radiomics software and the Image Biomarker Standardization Initiative provide useful definitions and reproducibility principles (Van Griethuysen et al., 2017; Zwanenburg et al., 2020), but their use does not automatically validate a color-histology pipeline.

**Table 2.** Stain specific texture composites.

| Composite | Standardized components |
| --- | --- |
| CD8 texture | Negative entropy, energy, coarseness |
| CD163 texture | Entropy, zone variance, busyness |
| H&E texture | Entropy, contrast, busyness |

Equal weighting gives each standardized component the same algebraic coefficient, but correlations between components can produce unequal effective contributions. The final mean composite is also not necessarily a unit-variance variable. Accordingly, a coefficient per composite unit should not be described as an effect per standard deviation unless the composite itself was standardized again. Stain- and scanner-related effects require separate assessment because reproducible numeric computation alone does not establish reproducible tissue measurement (Leo et al., 2016).

### 2.4 Survival modeling and performance interpretation

Clinical variables and stain-specific composites were entered into Cox proportional hazards models. Continuous texture features were standardized separately within each stain using z-scores calculated across patients before construction of the corresponding equal-weight composites. The Cox models included PSA, biopsy Gleason score, extraprostatic extension, seminal vesicle involvement, margin status, T stage, age, and the specified stain-specific composites. Models were fitted using the lifelines Cox proportional hazards implementation with an L2 penalizer of 0.1; no penalty-tuning procedure was performed. The model relates covariates to the instantaneous event hazard while leaving the baseline hazard unspecified. The linear predictor η and model hazard are given by the following expressions.

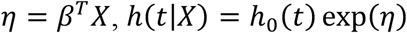

A coefficient β represents a conditional log-hazard association; its exponential is a hazard ratio for a one-unit change in the corresponding implemented predictor, with other covariates held constant (Cox, 1972). The risk score reported in this analysis was the partial hazard, exp(η), obtained using the model’s partial-hazard prediction, and therefore represents a relative hazard rather than an absolute probability of recurrence. The fitted model’s concordance index was used to describe discrimination. Proportional hazards assume constant covariate hazard ratios over time; no proportional-hazards test or patient-level influence diagnostic was performed in the present analysis (Grambsch & Therneau, 1994).

Four reported configurations were considered: clinical variables alone; clinical variables with H&E features; clinical variables with CD8 and CD163 features; and clinical variables with all three composites. Their reported C-indices are presented descriptively. In the absence of a documented patient-level evaluation procedure, these values must be interpreted as exploratory apparent discrimination, rather than estimates of performance in new patients (Harrell et al., 1996). Sample-size requirements depend on the number of estimated predictor parameters, event frequency, and anticipated model fit; a fixed events-per-variable rule does not establish adequacy (Riley et al., 2019).

The C-index assesses ordering of event times among evaluable pairs, with interpretation depending on the estimator and censoring (Uno et al., 2011). It does not measure probability calibration or the accuracy of a predicted recurrence date. Censoring-adjusted concordance, time-specific Brier scores, calibration, and decision-curve analysis address different aspects of model performance and clinical value(Gerds & Schumacher, 2006; Van Calster et al., 2019, 2019; Vickers & Elkin, 2006). These are requirements for further evaluation, not additional results established in this analysis.

### 2.5 Recurrence probabilities over time

For a fitted cumulative baseline hazard Hо, the Cox model produces a patient-specific event-free survival function. An interval probability measured from the original prediction time is the difference between survival probabilities at the interval boundaries.

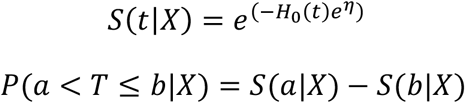

For a patient known to remain recurrence-free at the start of an interval, the corresponding conditional probability requires division by survival at that time, provided it is greater than zero.

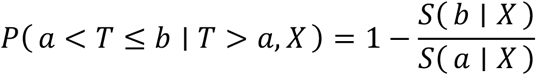

The unconditional quantity allocates baseline event probability across time windows. The conditional quantity answers the clinically different question of recurrence during the next interval among patients who have remained event-free to its start. Conditioning on survival alone does not update the model with new PSA measurements or treatments. Six-month windows were explored in the original implementation, and selecting the interval with the largest probability was treated as a descriptive display rather than a validated forecast of an individual’s recurrence date.

These survival-to-risk conversions assume the modeled event process is appropriate for the target population. If death before BCR is a competing event, simply censoring deaths and subtracting a cause-specific survival curve from one does not estimate the observed cumulative incidence of BCR. A competing-risk formulation is then needed for absolute-risk prediction (Fine & Gray, 1999). Likewise, selecting patients because recurrence was observed changes the population represented by the survival function.

## 3. Results

### 3.1 Cohort and measurement feasibility

The exploratory analysis included 15 patients with linked clinical and pathomic information. Three stain-specific patient-level composites were constructed and combined with clinical covariates, demonstrating the feasibility of reducing multiple tissue images to a compact input representation for survival modeling. The cohort consisted of patients who experienced BCR. The time to recurrence ranged from early events at approximately 4 months to extended follow-up up to 96 months, reflecting heterogeneity in disease progression.

### 3.2 Reported discrimination across model configurations

The clinical model had a reported C-index of 0.66, increasing to 0.68 with H&E features. A model incorporating CD8 and CD163 features was reported at approximately 0.88, while the model containing all three composites had a C-index of 0.89 (**Table 3**). These descriptive values suggest a signal worth investigating for immune-stain measurements, but the small sample and undocumented validation procedure preclude a reliable estimate of incremental performance.

**Table 3.** Reported discrimination by model configuration.

| Model | Reported C index | 95% CI |
| --- | --- | --- |
| Clinical variables | 0.66 | 0.38- 0.85 |
| Clinical variables and H&E | 0.68 | 0.38 - 0.86 |
| Clinical variables and CD8 and CD163 | 0.88 | 0.75-0.95 |
| Clinical variables and all three composites | 0.89 | 0.76- 0.96 |

### 3.3 Coefficient patterns

Approximate fitted coefficients were positive for the CD8 texture composite (β ≈ 1.06) and CD163 texture composite (β ≈ 0.51), and close to zero for the H&E texture composite (β ≈ −0.10). These values describe fitted conditional associations in this small dataset. Without their uncertainty, precise scaling, and resampling stability, they cannot establish independent prognostic effects or rank biological importance.

In the fitted models, PSA and extraprostatic extension had positive coefficient directions, whereas seminal vesicle invasion had a negative direction. The latter should not be interpreted as a protective biological effect. Sparse categories, correlated covariates, coding choices, and influential patients can change coefficient signs, particularly when stage and its component pathology variables are entered together. The available model specification does not support attributing these patterns to fitted interactions.

### 3.4 Risk-score modeling

Figure 2. shows an overall inverse association between model-derived risk scores and observed time to biochemical recurrence. Higher risk scores generally corresponded to earlier recurrence, whereas lower scores were associated with longer intervals to recurrence. This pattern is consistent with the interpretation of the Cox model, in which higher risk scores indicate a higher modeled hazard of recurrence. Outliers are noticed in a few cases where the risk score is high, but observed time to recurrence is longer. This discordant observation indicates an imperfect agreement between risk scores and recurrence timing within the analysis cohort. Their underlying causes cannot be determined from the present analysis. Evaluation in a larger, independent cohort is needed to assess the stability of the risk estimates and the reproducibility of the observed relationship.

### 3.5 Survival Analysis

Figure 3. presents patient-specific recurrence-free survival curves estimated using the Cox proportional hazards model. Higher risk scores generally corresponded to more rapid declines in estimated recurrence-free survival, whereas patients with lower scores maintained higher probabilities of remaining recurrence-free over time. These curves complement the relative risk scores by expressing the model’s estimates of recurrence-free survival across the evaluated time horizon.

Recurrence probabilities were subsequently calculated for predefined time intervals using the estimated survival functions. For each patient, the interval with the highest estimated recurrence probability was designated as the model-predicted recurrence window. Agreement between these windows and the observed recurrence intervals varied across patients. This interval-based approach provides an exploratory summary of the model’s estimated recurrence probability over time. Further evaluation of calibration and recurrence-window accuracy is needed to establish its reliability for individual prediction.

**Figure 2.**
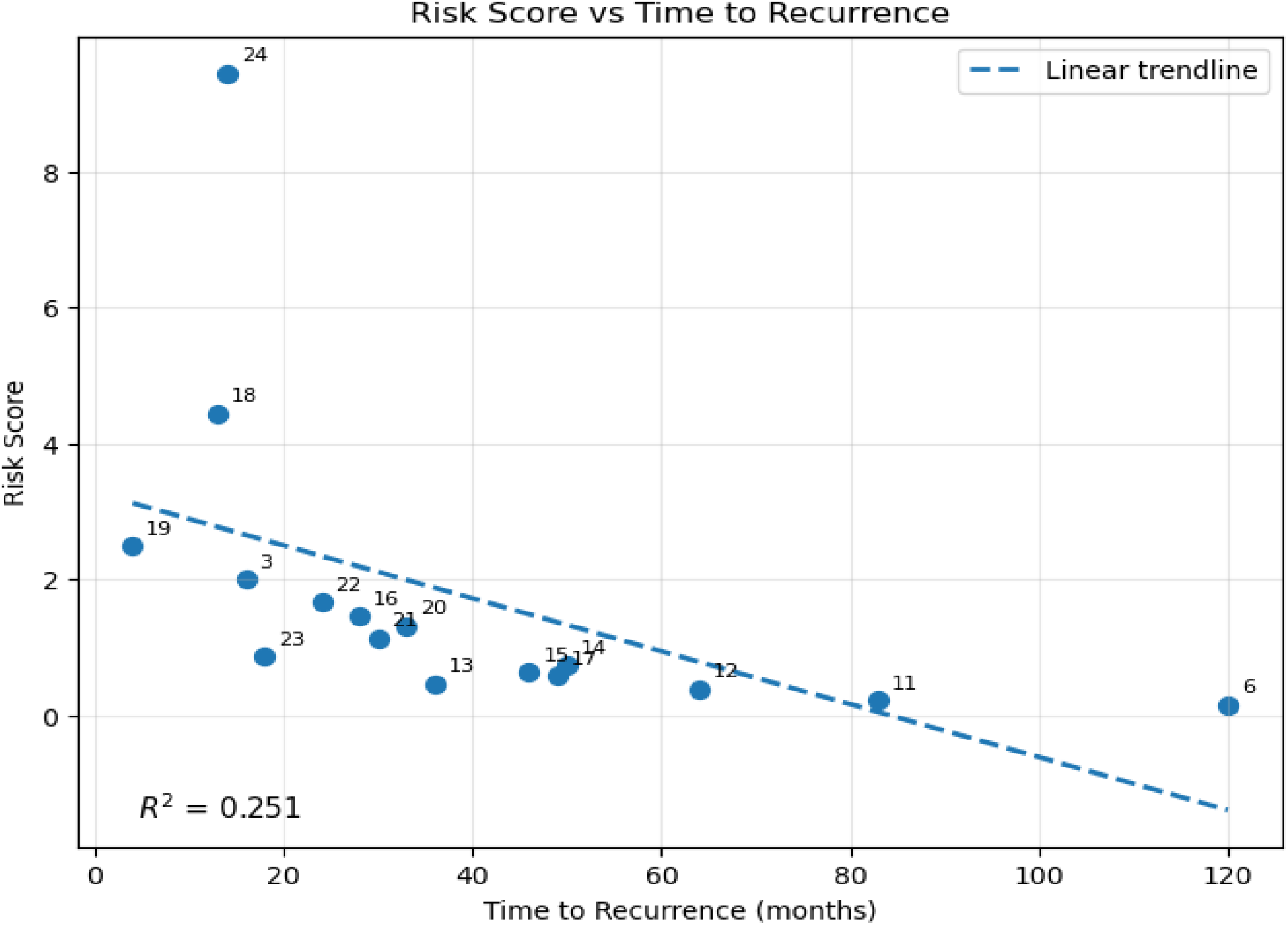
Relative hazard scores and recorded time. The scatterplot displays the overall inverse relationship between fitted relative hazard and observed time. The dashed line is a descriptive linear fit rather than a Cox-model prediction; its displayed R^2^ does not measure Cox-model discrimination or calibration

**Figure 3.**
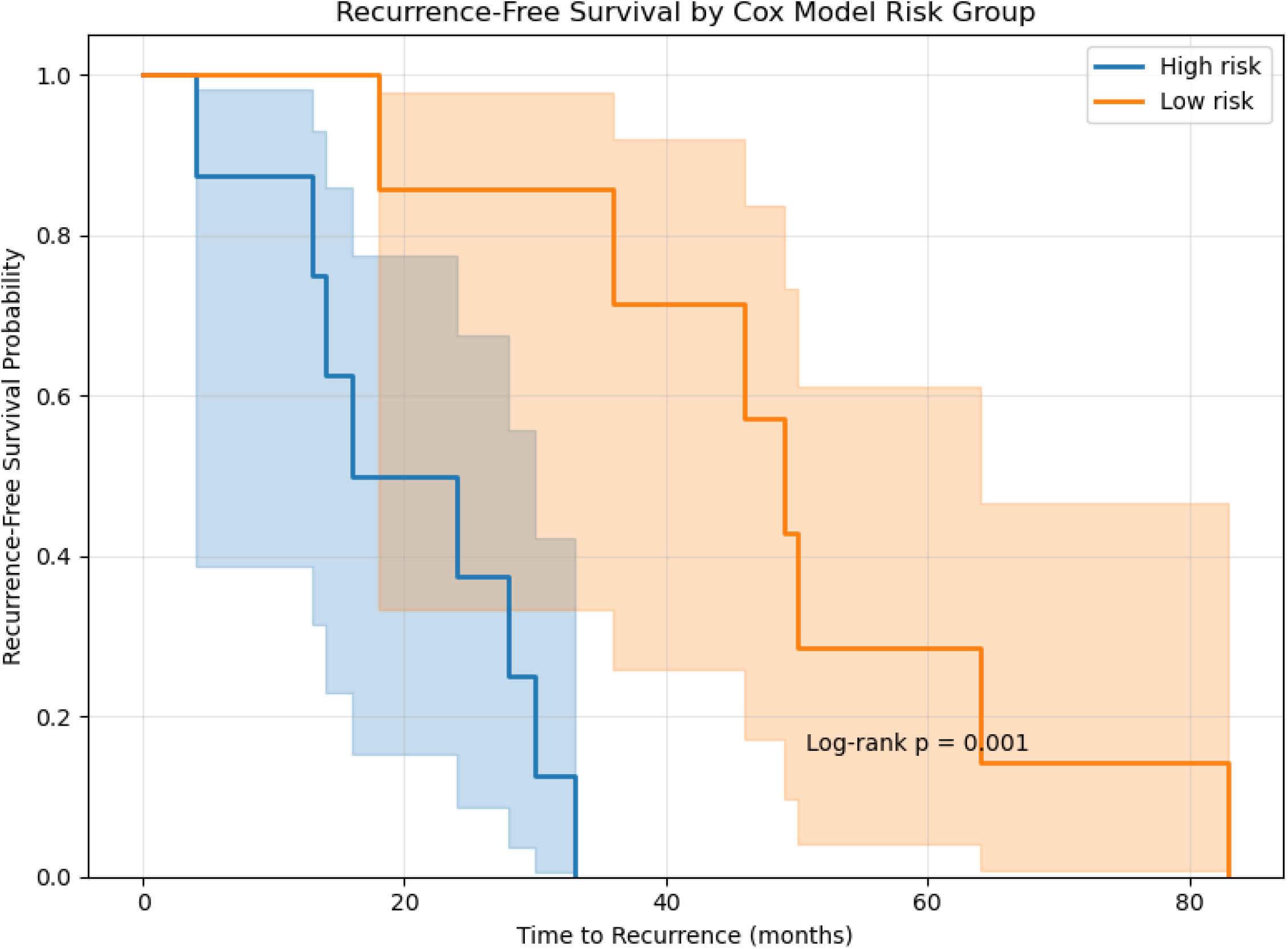
Exploratory recurrence-free survival by fitted risk group. Higher- and lower-risk groups are displayed with shaded uncertainty bands. This grouped display is distinct from an individual patient-specific Cox survival curve. Separation of groups constructed from the fitted model is an in-sample description rather than independent validation.

## 4. Discussion

In this exploratory cohort, adding the CD8 and CD163 texture composites to clinicopathologic predictors raised the apparent C-index from 0.66 to 0.88, whereas the H&E composite contributed little (0.68 with clinicopathologic variables; 0.89 with all three composites). The CD8 composite had the largest coefficient. These results suggest that the texture of immune staining, quantified without cell segmentation, may carry information about the timing of postoperative BCR that routine clinicopathologic variables do not capture. Given the cohort size, we regard this as a hypothesis to be tested rather than an estimate of the increment that would be observed in practice.

The size of the increment requires careful interpretation. When candidate predictors are numerous relative to events, adding variables raises apparent concordance even if they carry no information, and light L2 penalization only partly offsets this optimism. (Leo et al., 2021; Marin et al., 2025) These studies support the rationale for computational pathology but do not directly validate the present composites. The small, fitted H&E coefficient should be interpreted in relation to the selected texture features and model specification, rather than as evidence that prostate morphology lacks prognostic value.

The positive CD8 and CD163 coefficients require measurement-specific biological interpretation. Greater CD8-positive lymphocyte density, particularly within tumor epithelium, has been associated with shorter biochemical failure-free survival (Ness et al., 2014). Infiltrating regulatory T cells and macrophages have been associated with adverse biochemical recurrence outcomes, while high infiltration of peritumoral CD163-positive macrophages has been associated with late adverse outcomes (Andersen et al., 2021; Molina et al., 2023). The stain-specific composites developed in this study integrate established texture features extracted using PyRadiomics to summarize heterogeneity in CD8 and CD163 staining. Their associations with recurrence motivate further investigation of the prognostic information captured by these tissue patterns. Correlation with compartment-specific immune-cell density and functional markers would clarify the biological characteristics represented by the composites, including their relationships to T-cell activity and macrophage phenotypes.

A practical advantage of the automated quantification of tissue heterogeneity, including potentially subvisual patterns that are difficult to assess consistently through routine microscopy. PyRadiomics provides mathematically defined features describing intensity distributions and spatial relationships within selected image regions (Van Griethuysen et al., 2017). In our workflow, these features are extracted from preprocessed tissue images without requiring explicit identification and quantification of individual cells, epithelium, stroma, or lumina. This reduces reliance on detailed manual annotations or dedicated AI segmentation models. The stain-specific composites developed in this study further condense these measurements into a compact representation suitable for integration with clinicopathologic variables.

By characterizing staining heterogeneity and local spatial relationships, the proposed texture composites may provide prognostic information that is not fully represented by cell counts or compartment proportions. This offers the potential to improve recurrence prediction while reducing reliance on detailed cellular or compartmental segmentation. Whether these advantages translate into superior predictive performance should be evaluated through direct comparisons with models incorporating cell counts and tissue proportions in independent cohorts.

This cohort provides a focused setting for exploring how clinicopathologic variables and stain-specific texture composites relate to earlier versus later BCR after radical prostatectomy. The analysis generates hypotheses about recurrence timing among patients with observed BCR. Evaluation in larger, representative postoperative cohorts, including patients who remain recurrence-free, would clarify the generalizability of these associations and their value for estimating absolute recurrence probabilities (Wolff et al., 2019).

Model-derived survival curves and recurrence windows provide an intuitive temporal interpretation of the fitted risk score, with potential relevance to postoperative surveillance research. Further evaluation should distinguish predictions made at radical prostatectomy from those conditional on remaining recurrence-free and assess discrimination, calibration, and prediction error at clinically relevant horizons (Gerds & Schumacher, 2006; Van Calster et al., 2019; Uno et al., 2011).

The potential clinical contribution of this exploratory study is to refine the characterization of recurrence timing after radical prostatectomy using quantitative information derived from the surgical specimen. Clinicopathologic variables summarize established adverse features, while the stain-specific composites provide an additional description of morphological and immune-staining heterogeneity. The observed discrimination pattern supports investigating whether these measurements help distinguish earlier from later BCR among patients with otherwise similar postoperative findings. If validated in representative cohorts, this information could support studies of individualized PSA surveillance, postoperative risk communication, and timely multidisciplinary reassessment when recurrence is suspected. These applications provide a direct connection to salvage radiotherapy through improved postoperative prognostic assessment. Establishing their practical value requires demonstrating that the composites add reliable information beyond conventional predictors in cohorts that include patients who remain recurrence-free.

Several limitations warrant consideration. The small sample size relative to model complexity increases susceptibility to overfitting and coefficient instability, limiting confidence in the generalizability of the reported performance. Because the cohort comprised patients with observed BCR, the findings primarily describe recurrence timing within this group; their applicability to absolute recurrence risk in an unselected postoperative population remains to be established. Patient-level aggregation provides a compact representation of tissue features but may obscure localized morphological and immune-staining patterns. Evaluation in a single cohort without external validation further limits conclusions about performance across institutions and imaging conditions.

Nevertheless, this study demonstrates the feasibility of combining stain-specific texture composites with clinicopathologic variables to investigate time to BCR after radical prostatectomy. Model-derived survival curves and interval probabilities provide a temporal interpretation of the fitted risk scores and specific outputs for subsequent validation. Future studies should prioritize larger, representative, multi-institutional cohorts that include recurrence-free patients, consistent image preprocessing and composite construction, and independent assessment of discrimination, calibration, and prediction error with appropriate uncertainty estimates. Larger datasets could also support investigation of complementary molecular and imaging biomarkers and whether modeling nonlinear associations provides additional prognostic value.

## 5. Conclusion

Stain-specific pathomic texture composites offer a compact approach to integrating histologic and immune-staining heterogeneity with clinicopathologic predictors of BCR timing after radical prostatectomy. The observed increase in apparent discrimination with CD8 and CD163 composites supports further investigation of their incremental prognostic information. Establishing clinical value will require reproducible tissue measurements, adequately sized cohorts, and independent validation of both risk ranking and absolute recurrence probabilities.

## Data Availability

All data produced in the present study may be available upon reasonable request to the authors

